# Neighborhood Socioeconomic Status and Cardiometabolic Outcomes in Urban Jamaica: Exploring Novel Measures

**DOI:** 10.1101/2025.02.07.25321848

**Authors:** Harika P. Dyer, Trevor S. Ferguson, Joette Mckenzie, Novie Younger- Coleman, Marshall Tulloch-Reid, Alphanso L Blake, Nadia R. Bennett, Shelly McFarlane, Rainford J. Wilks, Tiffany L. Gary-Webb

**Affiliations:** Department of Epidemiology, University of Pittsburgh, School of Public Health, 130 DeSoto Street, Pittsburgh, PA 15261, United States; Epidemiology Research Unit, Caribbean Institute for Health Research Institute, The University of the West Indies, Mona, Kingston 7, Jamaica

**Keywords:** Chronic Disease, Cardiovascular Diseases, Social Determinants of Health, Socioeconomic Factors, Health Status Disparities, Vulnerable Populations, Jamaica, West Indies

## Abstract

**Introduction:** Cardiometabolic outcomes burden Afro-Caribbean populations disproportionately and their relationship with neighborhood socioeconomic status (SES) is unclear. This study explores neighborhood SES and cardiometabolic outcomes in urban Jamaica.

**Methods:** We analyzed data from 833 participants (women=557, men=276) to examine associations between neighborhood SES, determined by median property sales price, and measured diabetes, hypertension, high cholesterol, and obesity outcomes. We adjusted for covariates using survey-weighted Poisson regression models.

**Results:** High neighborhood SES was associated with increased prevalence of hypertension (PR=1.33, p=0.044) and high cholesterol (PR=2.46, p<0.001) in fully adjusted models compared with low neighborhood SES. We observed increased obesity for the middle (PR=1.46, p<0.001) and high (PR=1.60, p<0.001) SES tiers and several sex-specific relationships. High neighborhood SES was associated with more obesity among women (P=1.32, p=0.015) and high total cholesterol among men (PR=4.70, p=0.005). Mid SES was associated with increased hypertension among men (PR=2.02, p=0.024).

**Conclusion:** Associations between neighborhood SES and cardiometabolic outcomes in urban Jamaica appear to be nonlinear and influenced by a combination of sex, individual risk factors, and neighborhood characteristics. These findings challenge some conventional assumptions about the protective effects of higher SES on health and underscore the need for a robust understanding of the complex interplay individual and neighborhood determinants in similar contexts worldwide.

## Introduction

Diabetes, hypertension, high cholesterol, and obesity pose significant public health challenges in the Caribbean, where Afro-Caribbean populations face a globally disproportionate burden of these cardiometabolic outcomes.^1,2^ Socioeconomic status (SES), both at the individual and neighborhood level, is a recognized determinant of cardiometabolic health.^3,4^ Yet research exploring SES and cardiometabolic outcomes among Afro Caribbean populations has been limited, with evidence of nonlinear associations that differ for men and women.^5–8^ A positive association between obesity and family income was observed among Afro-Caribbean men in the United States but not among women, consistent with findings from Jamaica.^9,10^ In addition, high income has been linked to increased odds of metabolic syndrome among Jamaican men,^5^ while lower SES based on household possessions has been associated with higher odds of hypertension among young Jamaican women.^11^

At the neighborhood level, research findings are also mixed. Analyses of 2000-2008 data found that higher neighborhood SES was associated with lower blood pressure among Jamaican men, while poorer neighborhood infrastructure led to overweight and obesity among Jamaican women.^12,13^ However, a more recent study found that men residing in low-SES communities based on median land value had lower odds of ideal cardiovascular health compared to men living in high-SES communities, though the reverse was observed among women.^7^ A nonlinear association was observed regarding neighborhood disorder in urban Jamaica in another recent study.^6^

While these studies provide evidence that neighborhood SES plays a significant role in cardiometabolic outcomes in Jamaica and possibly the wider Caribbean, the nuances of the relationship and reasons for mixed results remain unclear. The present study seeks to add to the literature by examining the association between neighborhood SES—measured by median property sales price—and cardiometabolic outcomes, while accounting for other neighborhood SES characteristics such as average employment rates, education levels, and ratio of dependents to working-age population. Property price is a novel measure of community wealth and strongly predictive of health outcomes.^14,15^ We hypothesized that adjusting for other neighborhood SES variables will result in a stronger association between lower property prices and poorer cardiometabolic health.

## Methods

### Data Collection and Sample

#### Survey Data Collection

We conducted a cross-sectional analysis using demographic, health, and anthropometric data collected in the Cardiovascular Health in Urban Communities (CHUC) study. Details of the CHUC study design have been previously published.^16^ Trained interviewers administered questionnaires on demographic characteristics, medical history, cigarette smoking, physical activity, and dietary practices. Anthropometric measurements, blood pressure, fasting glucose, and total cholesterol were collected using standardized protocols.^7^

CHUC study participants (n = 849) were aged 15 years and older from the urban areas of four parishes constituting Jamaica’s Southeast Health Region (Kingston, St. Andrew, St. Catherine, and St. Thomas). These areas, classified as enumeration districts (EDs) by the Statistical Institute of Jamaica (STATIN), are the smallest geographic unit in Jamaica’s national household census, each comprising approximately 150 dwellings in urban areas.^17^ These data were used to group participants into communities (neighborhoods) with geographic bounds defined by the Social Development Commission (SDC) (https://sdc.gov.jm/about-us), the government agency overseeing Jamaica’s 775 communities.

Data was collected June 1, 2018 - June 30, 2019 and participants provided written informed consent and the study was approved by the University of the West Indies (UWI) Mona Campus Research Ethics Committee (MCREC) (ECP89 16/17). All data were deidentified by UWI researchers before they were accessed by researchers from the University of Pittsburgh.

#### Obtaining Property Sales Data

Property value is an emerging measure of socioeconomic status that is strongly predictive of health and has proven meaningful in studies of neighborhood SES and cardiometabolic outcomes,^14,15,18^ including a study of ideal cardiovascular health in Jamaica.^7^ Since property values include the value of the land and added improvements,^15^ we chose property sales prices as the primary neighborhood SES measure rather than unimproved property values.

We merged property sales data from Jamaica’s National Land Agency (NLA) JAMPROP tool (https://jampropsales.nla.gov.jm/about.aspx) with unimproved property data obtained for the CHUC study to match each property with its corresponding ED. The process for obtaining unimproved property data and overlaying EDs via shapefile has been previously published.^7^ The final dataset included all properties with sales data from 2003 to 2022 for 43 EDs. While the CHUC study included 44 EDs, there was no sales data for a Downtown Kingston community where eight CHUC study participants resided. The final dataset covered 1,083 parcels of land across 36 neighborhoods in the four parishes.

#### Obtaining Additional Neighborhood Characteristic Data

Additional socioeconomic characteristics for each neighborhood were obtained from community profiles purchased from the SDC: dependency ratio (ratio of dependents to working-age population), proportion of households receiving state assistance, and employment and educational attainment of household heads. SDC profiles were compiled by SDC Community Development Officers and published between 2004 and 2017.

#### Study Sample

Of 1130 participants targeted for the study, 849 consented to participate, for an overall response rate of 75%. Sixteen participants were not included in the analysis because their community data was missing. χ2 tests showed no significant differences in characteristics or outcomes when comparing these 16 participants to the remaining dataset. The final analytical sample consisted of 833 participants (557 women and 276 men).

### Measures

#### Neighborhood SES Exposure Variable

We selected median property sales price (including residential and commercial properties) as our primary exposure variable. Sales prices were converted to 2019 equivalent Jamaican dollars (JMD) to account for inflation.^19^ The most recent transaction was used for properties with multiple transactions. Median property sales price was calculated for each parish, ED, and neighborhood, with neighborhood SES represented by the neighborhood median. Neighborhoods were categorized into low SES (JMD 9,284 to 2.22M), mid SES (JMD 2.60M to 7.89M), and high SES (JMD 8.30M to 3.85B).

#### Covariates

The remaining neighborhood variables were selected as neighborhood-level covariates. Dependency ratio was defined as the number of dependents aged 0 to 14 and older than 65 divided by the neighborhood population aged 15 to 64, expressed as a percentage. A high dependency ratio indicates a high economic burden on the working-age population.^20^ Educational attainment represented the percentage of household heads with at least a secondary level education. Percentage of household heads employed and percentage of households receiving state assistance were also considered. For each variable, neighborhoods were classified as low, mid, and high SES, as reported in Table 1.

**Table 1.** Sociodemographic, cardiometabolic, and individual and neighborhood characteristics of urban Jamaicans ≥ 15 years in the Southeast Health Region stratified by sex.

|  | **Women**<br>**(n=557)** | **Men**<br>**(n=276)** | **Total**<br>**(n=833)** | **P-value<sup>1</sup>** |
| --- | --- | --- | --- | --- |
|  | n (%) | n (%) | n (%) |  |
| Age group |  |  |  | 0.191 |
| *Age <25 years* | 68 (12.2) | 48 (17.4) | 116 (3.9) |  |
| *Age 25 to 44 years* | 171 (30.7) | 79 (28.6) | 250 (30.0) |  |
| *Age 45 to 64 years* | 202 (36.3) | 89 (32.2) | 291 (34.9) |  |
| *Age $\geq 65$ years* | 116 (20.8) | 60 (21.7) | 176 (21.1) |  |
| Educational attainment |  |  |  | 0.921 |
| *Less than high school* | 124 (22.6) | 59 (21.4) | 183 (22.2) |  |
| *High school* | 285 (52.0) | 144 (52.4) | 429 (52.1) |  |
| *More than high school* | 139 (25.4) | 72 (26.2) | 211 (25.6) |  |
| Adequate physical activity ($\geq 150$ min moderate or 75 minutes of vigorous activity / week) | 232 (42.0) | 144 (52.4) | 384 (45.7) | 0.003* |
| Healthy diet ($\geq 3$ of 4 dietary factors)<sup>2</sup> | 133 (23.9) | 60 (21.7) | 193 (23.2) | 0.491 |
| Current smoker | 60 (10.8) | 82 (29.7) | 142 (17.0) | <0.001* |
| Measured or self-reported doctor diagnosis of diabetes | 101 (18.2) | 40 (14.5) | 141 (16.9) | 0.184 |
| Measured or self-reported doctor diagnosis of hypertension | 295 (53.0) | 126 (45.6) | 421 (50.5) | 0.047* |
| Measured or self-reported doctor diagnosis of high cholesterol | 191 (34.3) | 60 (21.7) | 255 (30.1) | <0.001* |
| Obesity (BMI $\geq 30$) | 245 (45.7) | 51 (19.4) | 296 (37.0) | <0.001* |
| Neighborhood SES (Median property sales price)<sup>3</sup> |  |  |  |  |
| *Low (JMD 9,284 to 2.22M)* | 198 (35.5) | 95 (34.4) | 293 (35.2) | 0.052** |
| *Mid (JMD 2.60M to 7.89M)* | 194 (34.8) | 78 (28.3) | 272 (32.6) |  |
| *High (JMD 8.30M to 3.85B)* | 165 (29.6) | 103 (37.3) | 268 (32.2) |  |
| Household heads with ≥ secondary education | | | | |
| *Low (17.9 to 42%)* | 165 (33.6) | 95 (39.4) | 260 (35.5) | 0.298 |
| *Mid (42.3 to 54.2%)* | 160 (32.6) | 73 (30.3) | 233 (31.8) |  |
| *High (54.5 to 62.3%)* | 166 (33.8) | 73 (30.3) | 239 (32.6) |  |
| Dependency ratio<sup>4</sup> |  |  |  |  |
| *Low (36.8 to 48.0%)* | 157 (31.7) | 92 (38.2) | 249 (33.8) | 0.196 |
| *Mid (48.8 to 57.0%)* | 171 (34.5) | 79 (32.8) | 250 (34.0) |  |
| *High (58.0 to 78.3%)* | 167 (33.7) | 70 (29.0) | 237 (32.2) |  |
| Household heads employed |  |  |  |  |
| *Low (36.4 to 58.8%)* | 157 (33.8) | 98 (42.6) | 255 (36.7) | 0.057** |
| *Mid (59.0 to 70.0%)* | 194 (41.7) | 78 (33.9) | 272 (39.1) |  |
| *High (70.5 to 75.0%)* | 114 (24.5) | 54 (23.5) | 168 (24.17) |  |
| Households receiving state assistance |  |  |  |  |
| *Low (3.6 to 8.6%)* | 207 (46.6) | 81 (37.7) | 288 (43.7) | 0.073** |
| *Mid (9.0 to 14.8%)* | 117 (26.3) | 61 (28.4) | 178 (27.0) |  |
| *High (16.0 to 24.3%)* | 120 (27.0) | 73 (33.9) | 193 (29.3) |  |

\*Statistically significant differences between women and men (p < 0.05)
\*\*Marginally significant differences between women and men (0.05 < p < 0.075)
1. P-values were calculated using $\chi^2$ tests.
2. Participants were classified as having a healthy diet if they $\geq 3$ of the following dietary characteristics: (1) low-salt diet, defined as no added salt at the table and rarely or never eats processed foods; (2) low sugar-sweetened beverage (SSB) consumption, defined as SSB intake less than twice per week; (3) adequate fruit and vegetable consumption, defined as intake of fruits or vegetables $\geq 3$ times per day; and (4) adequate fish consumption, defined as eating fish $\geq 2$ times per week.
3. Median property sales prices at the neighborhood level, with neighborhoods categorized as low, mid, and high. Reported in Jamaican dollars (JMD).
4. Dependency ratio is the number of dependents age 0 to 14 and over the age of 65, compared with the total population age 15 to 64, expressed as a percentage.

Individual characteristics were included as covariates. Age was categorized into < 25 years, 25 to 44 years, 45 to 64 years, and ≥ 65 years. Sex was classified as female or male. Individual educational attainment was classified as “less than high school” for participants educated up to grade 8, “high school” for participants with some secondary education, and “more than high school” for participants educated beyond the secondary level. The remaining covariates were coded as binary variables. Participants who reported never smoking or having quit for longer than 12 months were classified as non-smokers. Participants engaging in adequate physical activity were those who were moderately active for at least 150 minutes each week or vigorously active for at least 75 minutes each week.^21^ Healthy diet was coded for participants who reported meeting three or more of the following criteria: (1) low-salt diet, defined as no added salt at the table and “rarely” or “never” eating processed foods; (2) low sugar-sweetened beverage (SSB) consumption, defined as consuming fewer than two SSBs each week; (3) adequate fruit and vegetable consumption, defined as eating fruits or vegetables three or more times per day; and (4) adequate fish consumption, defined as eating fish two or more times each week.^22^ Dietary fiber was not included as the CHUC study did not obtain whole grain intake data.

#### Cardiometabolic Outcome Variables

Cardiometabolic outcomes were categorized based on the Jamaican context and clinical standards at the time of data collection. Diabetes was defined as fasting glucose ≥ 7.0 mmol/L or a self-reported physician diagnosis of diabetes^23^ and hypertension as an average systolic blood pressure ≥ 140 mmHg or average diastolic blood pressure ≥ 90 mmHg for the second and third of three consecutive blood pressure readings, or a self-reported physician diagnosis of hypertension.^24^ High cholesterol was defined as total cholesterol ≥ 5.2 mmol/L or a self-reported physician diagnosis of high cholesterol.^25^ Obesity was defined as BMI ≥ 30 kg/m^2^.^26^

#### Statistical Analysis

Analyses were performed using Stata V.17.0. Crude and sex-specific descriptive statistics were produced for outcomes, exposures, and covariates. There was no statistically significant interaction between sex and the primary exposure variable in its relationship to the outcomes. However, we made an a priori decision to stratify by sex because of previous work in Jamaica showing variations in cardiometabolic outcomes and their relationship to exposures for men and women.^5–8^ Differences in characteristics between men and women and across neighborhood SES categories were compared using χ^2^ tests. The prevalence of each outcome was also estimated across sex and neighborhood SES using three weighted Poisson regression models. Weights accounted for the survey sampling design and the age and sex distribution of the Jamaican population. As a sensitivity analysis, multilevel models found that clustering did not contribute to variability in the relationship between the primary exposure variable and the outcomes. Model 1 estimated crude prevalence ratios (PR). Model 2 adjusted for individual risk factors (sex, age group, education level, smoking, diet, and physical activity). Model 3 adjusted for individual risk factors and neighborhood covariates (education level, dependency ratio, employment, and state assistance). Weighted Poisson is an established method for estimating prevalence ratios.^27^ Equality of mean and variance was assessed for all models and variance inflation factor was assessed for adjusted models to satisfy regression assumptions. A listwise deletion approach was applied to missing data.

## Results

### Sample Characteristics

Table 1 summarizes participant characteristics. Most participants were aged 45 to 64 (34.9%) and had at least a high school education (77.7%). Hypertension was the most prevalent cardiometabolic outcome (50.5%), followed by obesity (37.0%). High cholesterol was present in 30.1% of participants, while 16.9% had diabetes.

Men were more likely to be adequately physically active (52.4% vs. 42.0%, p = 0.003) and current smokers (29.7% vs. 10.8%, p <0.001) compared to women. Women had higher rates of hypertension (53.0% vs. 45.6%, p = 0.047), high cholesterol (34.3% vs. 21.7%, p <0.001), and obesity (45.7% vs. 19.4%, p <0.001). There were marginal differences between men and women in the distribution of neighborhood SES (p = 0.052) and average proportion of household heads employed (p = 0.057).

### Neighborhood SES Characteristics

High SES neighborhoods had an average property sales price of JMD 20.07 million (n = 13, SD = JMD 10.07 million), compared to JMD 1.90 million (n = 12, SD = JMD 1.10 million) for low SES neighborhoods and 5.97 million (n = 11, SD = JMD 1.32 million) for mid SES. Figure 1 displays the distribution of median property sales prices by neighborhood SES.

**Figure 1.**
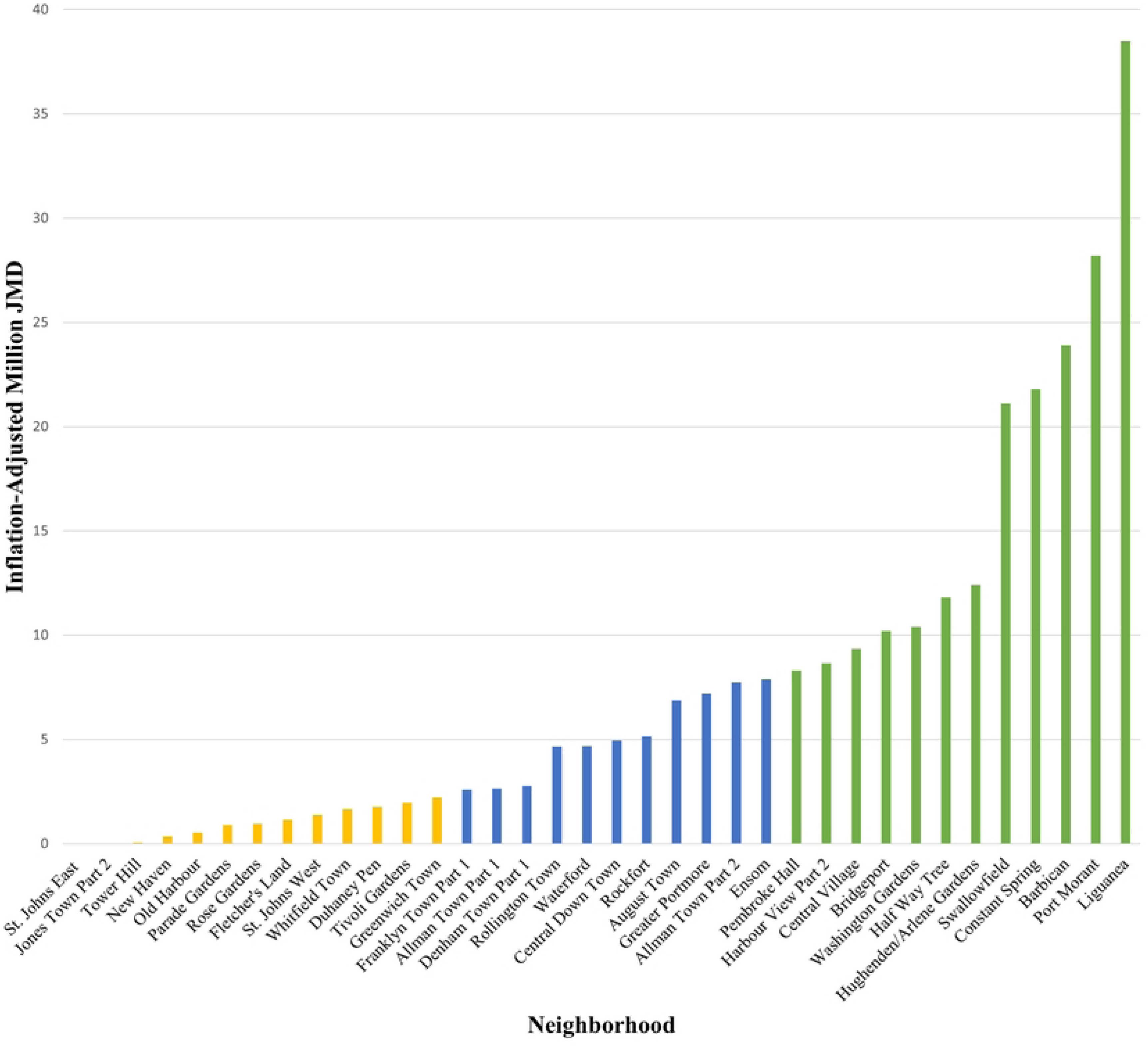
Median Property Sales Price and SES Categories Across Urban Neighborhoods in Jamaica’s Southeast Health Region.

### Neighborhood-Level Covariates

High cholesterol was more common among participants living in neighborhoods with low economic burdens, as indicated by low dependency ratios, versus those in neighborhoods with high dependency ratios (36.5% vs. 24.5%, p = 0.004). In neighborhoods with low dependency ratios, marginally more women and significantly more men had high cholesterol compared to areas with mid-level dependency ratios (women: 43.0% vs. 33.0%, p = 0.050; men: 34.0% vs. 17.9%, p <0.016).

Participants in neighborhoods with high household head employment had lower rates of diabetes compared to those in low (10.8% vs. 20.8%, p = 0.007) and mid-level employment neighborhoods (10.8% vs. 18.4%, p = 0.033). No significant associations were observed when men and women were considered separately.

In addition, household head education level and state assistance were not significantly linked to any of the cardiometabolic outcomes considered in this study.

### Individual-Level Covariates

The distributions of individual risk factors in relationship to SES as determined by median property price are shown in the Appendix. Participants in high SES neighborhoods were more likely to be 65 or older (28.0% vs. 18.4%, p = 0.039) and have education beyond high school (43.4% vs. 15.3%, p <0.001) in comparison to participants in low-SES neighborhoods. Age differences were significant for women, but not men, while education differences were significant for both in stratified analyses. Participants in the highest property price tier had healthier diets than those in the lowest tier (34.7% vs. 20.5%, p <0.001). This difference was significant for women only in stratified analyses. Participants in neighborhoods with high property prices were less likely to engage in adequate physical activity compared to those in neighborhoods with low property prices (42.0% vs. 53.1%, p = 0.009). Those in middle-tier neighborhoods also had lower levels of physical activity (41.1% vs. 53.1%, p = 0.005). In stratified analyses these differences were significant in women only, with no significant findings among men. Smoking was higher in neighborhoods with low property prices compared to those with high prices (21.8% vs. 9.33%, p <0.001). This difference was significant in men but not women in stratified comparisons.

### Regression Results

Tables 2 through 4 display prevalence ratios for each cardiometabolic outcome in relation to median property sales price, our primary neighborhood SES measure. Notably, high cholesterol and obesity exhibit the strongest associations with property prices.

**Table 2.** Survey-weighted prevalence ratios for association between neighborhood SES and cardiometabolic outcomes among urban Jamaican men and women ≥ 15 years in the Southeast Health Region.

|  | Diabetes |  | Hyper-tension |  | High Cholesterol |  | Obesity |  |
| --- | --- | --- | --- | --- | --- | --- | --- | --- |
|  | PR<br>(95% CI) | P-value | PR<br>(95% CI) | P-value | PR<br>(95% CI) | P-value | PR<br>(95% CI) | P-value |
| **Crude** |  |  |  |  |  |  |  |  |
| *Low (ref)* | – |  | – |  | – |  | – |  |
| *Mid* | 2.07<br>(1.18, 3.62) | 0.012 | 1.05<br>(0.82, 1.33) | 0.696 | 1.36<br>(0.93, 1.97) | 0.107 | 1.26<br>(0.98, 1.62) | 0.066 |
| *High* | 1.43<br>(0.70, 2.93) | 0.321 | 1.23<br>(1.03, 1.47) | 0.019 | 2.53<br>(1.78, 3.61) | <0.001 | 1.26<br>(1.00, 1.58) | 0.051 |
| **Adjusted for individual risk factors only<sup>1</sup>** |  |  |  |  |  |  |  |  |
| *Low* | – |  | – |  | – |  | – |  |
| *Mid* | 2.28<br>(1.38, 3.77) | 0.002 | 1.09<br>(0.89, 1.32) | 0.390 | 1.36<br>(0.97, 1.90) | 0.074 | 1.35<br>(1.09, 1.68) | 0.006 |
| *High* | 1.42<br>(0.71, 2.85) | 0.310 | 1.15<br>(0.92, 1.44) | 0.197 | 2.20<br>(1.48, 3.26) | <0.001 | 1.26<br>(0.98, 1.61) | 0.070 |
| **Adjusted for individual and neighborhood risk factors<sup>2</sup>** |  |  |  |  |  |  |  |  |
| *Low* | – |  | – |  | – |  | – |  |
| *Mid* | 0.81<br>(0.47, 1.39) | 0.426 | 1.08<br>(0.84, 1.38) | 0.540 | 1.90<br>(0.60, 1.74) | 0.924 | 1.46<br>(1.20, 1.78) | 0.001 |
| *High* | 0.68<br>(0.30, 1.54) | 0.347 | 1.33<br>(1.01, 1.74) | 0.044 | 2.46<br>(1.57, 3.84) | <0.001 | 1.60<br>(1.22, 2.11) | 0.001 |

1. Adjusted for sex, age group, education level, smoking, diet, and physical activity.
2. Adjusted for individual factors, as well as neighborhood averages of head of household education level, dependency ratio, head of household employment, and household state assistance.

**Table 3.** Survey-weighted prevalence ratios for association between neighborhood SES and cardiometabolic outcomes among urban Jamaican women ≥ 15 years in the Southeast Health Region.

|  | **Diabetes** |  | **Hyper-tension** |  | **High Cholesterol** |  | **Obesity** |  |
| --- | --- | --- | --- | --- | --- | --- | --- | --- |
|  | **PR**<br>**(95% CI)** | **P-value** | **PR**<br>**(95% CI)** | **P-value** | **PR**<br>**(95% CI)** | **P-value** | **PR**<br>**(95% CI)** | **P-value** |
| **Crude** |  |  |  |  |  |  |  |  |
| *Low (ref)* | – |  | – |  | – |  | – |  |
| *Mid* | 1.52<br>(0.86, 2.70) | 0.146 | 0.83<br>(0.51, 1.36) | 0.447 | 1.78<br>(0.71, 1.95) | 0.514 | 1.29<br>(0.98, 1.69) | 0.067 |
| *High* | 1.49<br>(0.67, 3.32) | 0.324 | 1.26<br>(0.96, 1.66) | 0.093 | 2.04<br>(1.29, 3.23) | 0.003 | 1.14<br>(0.89, 1.46) | 0.280 |
| **Adjusted for individual risk factors only<sup>1</sup>** |  |  |  |  |  |  |  |  |
| *Low* | – |  | – |  | – |  | – |  |
| *Mid* | 1.75<br>(1.00, 3.08) | 0.051 | 0.85<br>(0.55, 1.30) | 0.443 | 1.13<br>(0.73, 1.75) | 0.560 | 1.35<br>(1.06, 1.73) | 0.017 |
| *High* | 1.03<br>(0.47, 2.26) | 0.934 | 1.03<br>(0.80, 1.32) | 0.819 | 1.45<br>(0.88, 2.39) | 0.138 | 1.09<br>(0.83, 1.43) | 0.510 |
| **Adjusted for individual and neighborhood risk factors<sup>2</sup>** |  |  |  |  |  |  |  |  |
| *Low* | – |  | – |  | – |  | – |  |
| *Mid* | 0.74<br>(0.36, 1.51) | 0.393 | 0.72<br>(0.44, 1.17) | 0.178 | 1.12<br>(0.56, 2.23) | 0.735 | 1.30<br>(1.00, 1.69) | 0.053 |
| *High* | 1.10<br>(0.58, 2.07) | 0.762 | 1.25<br>(0.95, 1.63) | 0.106 | 1.81<br>(0.96, 3.40) | 0.065 | 1.32<br>(1.06, 1.65) | 0.015 |

1. Adjusted for sex, age group, education level, smoking, diet, and physical activity.
2. Adjusted for individual factors, as well as neighborhood averages of head of household education level, dependency ratio, head of household employment, and household state assistance.

**Table 4.** Survey-weighted prevalence ratios for association between neighborhood SES and cardiometabolic outcomes among urban Jamaican men ≥ 15 years in the Southeast Health Region.

|  | **Diabetes** |  | **Hyper-**<br>**tension** |  | **High**<br>**Cholesterol** |  | **Obesity** |  |
| --- | --- | --- | --- | --- | --- | --- | --- | --- |
|  | **PR**<br>**(95% CI)** | **P-**<br>**value** | **PR**<br>**(95% CI)** | **P-**<br>**value** | **PR**<br>**(95% CI)** | **P-**<br>**value** | **PR**<br>**(95% CI)** | **P-**<br>**value** |
| **Crude** |  |  |  |  |  |  |  |  |
| *Low (ref)* | – |  | – |  | – |  | – |  |
| *Mid* | 2.70<br>(1.03, 7.07) | 0.043 | 1.34<br>(0.90, 2.00) | 0.143 | 1.87<br>(0.74, 4.73) | 0.177 | 1.46<br>(0.72, 2.95) | 0.282 |
| *High* | 1.39<br>(0.58, 3.34) | 0.446 | 1.24<br>(0.85, 1.82) | 0.254 | 3.83<br>(1.53, 9.57) | 0.005 | 2.13<br>(1.15, 3.94) | 0.018 |
| **Adjusted for individual risk factors only<sup>1</sup>** |  |  |  |  |  |  |  |  |
| *Low* | – |  | – |  | – |  | – |  |
| *Mid* | 2.28<br>(1.04, 6.94) | 0.042 | 1.40<br>(0.98, 2.00) | 0.062 | 1.90<br>(0.76, 4.72) | 0.161 | 1.52<br>(0.70, 3.29) | 0.276 |
| *High* | 1.73<br>(0.67, 4.43) | 0.248 | 1.33<br>(0.89, 1.99) | 0.162 | 3.97<br>(1.43, 10.97) | 0.009 | 1.83<br>(0.87, 3.86) | 0.107 |
| **Adjusted for individual and neighborhood risk factors<sup>2</sup>** |  |  |  |  |  |  |  |  |
| *Low* | – |  | – |  | – |  | – |  |
| *Mid* | 0.67<br>(0.22, 2.02) | 0.469 | 2.02<br>(1.10, 3.71) | 0.024 | 0.99<br>(0.27, 3.63) | 0.983 | 4.24<br>(0.69, 26.18) | 0.115 |
| *High* | 0.34<br>(0.08, 1.43) | 0.136 | 1.69<br>(0.83, 3.45) | 0.143 | 4.70<br>(1.64, 13.44) | 0.005 | 5.06<br>(0.74, 34.71) | 0.096 |

1. Adjusted for sex, age group, education level, smoking, diet, and physical activity.
2. Adjusted for individual factors, as well as neighborhood averages of head of household education level, dependency ratio, head of household employment, and household state assistance.

Residing in high SES neighborhoods was associated with greater high cholesterol prevalence compared to low SES neighborhoods (PR = 2.53, p <0.001). This association remains robust after adjusting for individual risk factors (PR = 2.20, p<0.001) and other neighborhood SES characteristics (PR = 2.46, p <0.001). When stratified by sex, men in high SES neighborhoods were more likely to have high cholesterol (PR = 3.83, p = 0.005), with this relationship persisting after adjusting for individual (PR = 3.97, p = 0.009) and neighborhood factors (PR = 4.70, p = 0.005). Women in high SES neighborhoods had greater high cholesterol prevalence (PR = 2.04, p = 0.003), but this association was not significant after adjusting for the effects of individual- and neighborhood-level factors.

Obesity had a marginal crude association with property price in the middle (PR = 1.26, p = 0.066) and high (PR = 1.26, p = 0.051) SES categories. Adjusting for individual risk factors strengthened this association for the middle SES tier (PR = 1.35, p = 0.006), although the link remained marginal for the high SES tier (PR = 1.26, p = 0.070). After adjusting for other neighborhood characteristics, both the middle and high SES tiers had increased obesity prevalence when compared to the low SES tier (middle PR = 1.46, p <0.001; high: PR = 1.60, p <0.001). These relationships remained somewhat similar in models assessing women only. Obesity prevalence was higher among women in the middle SES tier after adjusting for individual risk factors (PR = 1.35, p = 0.017). After accounting for neighborhood characteristics (PR =1.30, 0.053), the association was shown to be marginal. Women in high SES neighborhoods also experienced higher levels of obesity when controlling for individual and neighborhood covariates (PR = 1.32, p = 0.015), although significant associations were not observed in crude associations or when considering individual covariates alone. Conversely, among men, crude analyses indicated higher obesity prevalence among high SES residents (PR = 2.13, p = 0.018). This association did not remain significant after adjusting for differences in individual and neighborhood characteristics.

We observed a significant association between hypertension and median property price, particularly for the high SES category when compared to low SES (PR = 1.23, p = 0.019). This association disappeared when adjusting for individual risk factors only, but reemerged when neighborhood characteristics were added to the model (PR = 1.33, p = 0.044). However, when stratified by sex, significant associations were observed only among men, with the middle SES residents exhibiting increased hypertension compared to the low SES residents (PR = 2.02, p = 0.024).

Regarding diabetes, significant associations with neighborhood property prices were observed for the middle SES group. In models unstratified by sex, there was a higher diabetes prevalence among participants in middle-SES neighborhoods compared to low-SES ones (PR = 2.07, p = 0.012), which remained significant after adjusting for individual risk factors (PR = 2.28, p = 0.002). However, significance was not retained when neighborhood characteristics beyond property prices were considered. These patterns persisted among men, with higher diabetes prevalence observed for the middle SES tier only in crude models (PR = 2.70, p = 0.043) and individual risk factor adjustments (PR = 2.28, p = 0.042). No significant associations between diabetes and property prices were found among women, although there was a marginally significant increase in prevalence for the middle tier when individual risk factor characteristics were accounted for (PR = 1.75, p = 0.051).

## Discussion

### Prevalence of Cardiometabolic Outcomes—Our Sample in Context

Women were overrepresented in the sample compared to Jamaica population estimates (66.9% vs. 50.5%), and more than half of participants were aged 45 or older (56.0%), reflecting a higher proportion of older persons relative to the general population.^17^ Hypertension, the most prevalent outcome, was recorded in 50.5% of participants (women: 53.0%, men: 45.6%). This pattern aligns with previous findings in Jamaica^28^ and suggests a consistent disparity by sex. Obesity was twice as prevalent in women when compared with men in our sample (45.7% vs. 19.4%), consistent with previous research.^29^ Interestingly, diabetes was also higher among women. While this aligns with previously published analyses of urban Jamaica, it contrasts with global patterns where men typically have similar or higher diabetes prevalence.^30,31^

### Cardiometabolic Outcomes and Neighborhood SES

Few studies have explored the associations between neighborhood SES with cardiometabolic outcomes in Jamaica. Moreover, existing research has shown non-linear associations, with some studies indicating increased risk in middle or high SES tiers compared to low SES.^10,12^ The reasons for these non-linear relationships are not clear, and we hypothesized that adjusting for other neighborhood SES variables would result in a more linear association between lower property prices and poorer cardiometabolic health. In our study, however, high neighborhood SES was associated with an increased prevalence of hypertension (PR = 1.33, p = 0.044) and high cholesterol (PR = 2.46, p <0.001), and both high SES (PR = 1.60, p = <0.001) and middle SES (PR = 1.46, p <0.001) were associated with increased obesity when compared with the lowest SES tiers. In stratified analyses, high SES was associated with more obesity among women (PR = 1.32, p = 0.015) and more high total cholesterol among men (PR = 4.70, p = 0.005). Mid-tier SES was also associated with greater rates of hypertension among men (PR = 2.02, p = 0.024).

Our findings underscore the complexity of the relationship between neighborhood SES and cardiometabolic outcomes. They align with other studies that have observed increasing hypertension prevalence with income in educated men^32^ and higher obesity among urban middle class Jamaicans who do not reside near public parks.^33^ Analyses from the Jamaican Health and Lifestyle Survey 2017-2016 showed that living in lower-SES communities was associated with reduced odds of meeting at least five of seven ideal cardiovascular health criteria (ICH-5) among Jamaican men and found the reverse to be true for educational attainment. Yet for Jamaican women educational attainment was instead linked with improved ICH-5.

Jamaica’s status as an upper-middle income country undergoing an epidemiologic transition, coupled with uneven economic development within its urban centers, may contribute to these non-linear associations.^34,35^ Similar relationships between SES and cardiovascular disease risk factors have been noted in other countries experiencing similar transitions.^36^ Cardiovascular disease prevalence was also historically higher among people of higher SES in high income countries in the early 20th century, when smoking, unhealthy diets and sedentary lifestyles were more common among high-income groups.^37^ Less privileged Jamaicans may face an even greater burden of cardiometabolic outcomes and cardiovascular disease over time if the same patterns persist in the Caribbean.

### Strengths

A strength of this paper is its use of a population-based sampling approach and survey weighting of regression analyses to produce prevalence ratios that were generalizable to Jamaica’s urban population in the southeast region. This study also adjusted not only for individual risk factors, but also neighborhood SES characteristics that could affect the relationship between property prices and cardiovascular health, such as household employment and education levels, dependency ratios, and state assistance. The use of property sales prices was also novel and presents more up-to-date representations of value when compared to unimproved property values reported by the NLA.^7^

### Limitations

While using property sales data as our primary SES indicator resulted in more accurate depictions of property worth when compared to unimproved property values, a consequence of this approach is that properties not sold in the 2003 to 2022 period were not included in the analyses. Future research could consider incorporating both sales price data and unimproved property values. In addition, low-income neighborhoods where informal property ownership is more common may have had fewer property sales. Moreover, neighborhood SDC profiles from which household employment and education levels, dependency ratios, and state assistance data were derived for this study were published over a span of 14 years from 2004 to 2017. While this matches the timespan for property data included in our analyses, it also means that SDC data for some neighborhoods is much older than data in other neighborhoods, which may affect comparability. A third limitation is the unavailability of data about alcohol use and marital status, which may have effects at the individual and neighborhood level.^38^

## Conclusions

Among urban Jamaicans, associations between cardiometabolic outcomes and neighborhood SES appear to be nonlinear and influenced by sex, individual risk factors, and neighborhood level confounders. When we adjusted for individual risk factors and other neighborhood characteristics, we found that urban Jamaican women living in neighborhoods with higher average property prices had a 32% higher prevalence of obesity, while urban Jamaican men had 4.7 times the prevalence of high cholesterol compared to counterparts living in neighborhoods with low property prices. Men living in mid-tier property price neighborhoods had double the prevalence of hypertension in comparison to those in lower-priced neighborhoods. These findings align with Jamaica’s epidemiologic transition and trends observed in other low- and middle-income countries. Further research incorporating a wider variety of individual and neighborhood-level SES indicators may provide additional insights into these relationships. Understanding relationships between neighborhood SES and adverse cardiometabolic outcomes is crucial for reducing the disproportionate burden borne by Jamaica and other Afro-Caribbean populations.

## Data Availability

The data underlying the results presented in the study are available from the Caribbean Institute for Health Research Institute at The University of the West Indies, Mona, Kingston 7, Jamaica

## Acknowledgements

The authors declare no potential conflicts of interest with respect to the research, authorship, or publication of this article. The authors received funding from the Bernard Lown Scholars in Cardiovascular Health Program (Award Number BLSCHP-1604) and the University of Pittsburgh Year of Pitt Global. No copyrighted material, surveys, instruments, or tools were used in the research described in this article. This research uses deidentified data from a study approved by the University of the West Indies (UWI) Mona Campus Research Ethics Committee (MCREC) (ECP89 16/17) and was exempt from additional Institutional Review Board approval from the University of Pittsburgh. All authors participated in this work and have reviewed and approved this manuscript. This work has not been previously published elsewhere.

